# A pilot study to see any Change of the Nasal and Oropharyngeal Microbiota with Prolonged Use of Medical Masks during the COVID-19 Outbreak

**DOI:** 10.1101/2020.08.15.20175067

**Authors:** Sayak Roy, Pampi Majumder, Kingshuk Bhattacharya

## Abstract

**Background:** The outbreak of coronavirus disease 2019 (COVID-19) has played havoc on the healthcare system and society. Many international guidelines have put forward various measures to control the spread and, using various quality masks seems to be the most important amongst them. This was a cross-sectional pilot study to see any alterations in the bacterial flora of the nasal and the oropharyngeal (OP) microbiota with the use of medical masks over prolonged periods during this COVID-19 outbreak.

**Methods:** Nasal and oropharyngeal swabs were collected using proper international guidelines from 30 healthy healthcare workers matching pre-set inclusion criteria, who gave written informed consent. The swabs were used for gram stain as well as culture and sensitivity analysis using standard methods.

**Results:** In general, we found that the oropharyngeal microflora harboured a more diverse population of bacteria (n=13) than the nasal microflora (n=5). The predominant bacterial flora was found to *Staphylococcus epidermidis* in the nasal cavity and *Streptococcus viridans* in the oropharyngeal cavity. There was no growth in 8 (26.68%) samples of oropharynx and 3 (10%) of nasal samples, with one patient having no growth in both the samples. The commonest resistant antibiotic from both the cavity cultures was benzylpenicillin (nasal flora 80% and OP flora 47.37%).

**Conclusion:** This small pilot study has shown a reassuring aspect of no change in the typical bacterial microflora species of the nasal and OP cavity with prolonged use of medical masks. This is the first study to show this convincing evidence during the COVID-19 outbreak and also in healthy healthcare workers who have to wear masks over long durations.

## Introduction

Since its beginning in December 2019, the novel coronavirus disease (COVID-19) has reached a death toll of 580045 cases and affected 13 378 853 patients till July 16, 2020 [1]. Preventive measures have been regarded as the primary combating tool and, various bodies have recommended various strategies to prevent this disease since only no specific treatments are available to date. Use of Personal Protective Equipment (PPE), social distancing, sanitizers, using masks, and hand-washing are the main preventive strategies recommended in most of the international and national guidelines for all healthcare personals and general people [2]. While Centers for Disease Control and Prevention (CDC) recommends using masks or N95 respirators for both low and high-risk situations, we have to remember that in severely frail patients like those on hemodialysis, masks can cause a significant drop in PaO2 level (101.7 +/− 12.6 to 92.7 +/− 15.8 mm Hg, p = 0.006) [3,4]. Another prospective study of the use of masks in 44 healthy participants undergoing a six minutes walking test showed that the use of masks was associated with significant dyspnea variation (p< 0.001) [5].

The microbes residing inside our body are termed as microbiota, and they form an integral part of our commensal health [6].

There is a difference in microbiota constituents between adults and children, and there is also evidence of denser microflora load in children with less diversity than adults [7, 8]. Studies have also shown the presence of many nontuberculous mycobacteria like *Mycobacterium gordonae* (8%), *Mycobacterium kansasii* (2%), *Mycobacterium fortuitum* (3%) and *Mycobacterium avium complex* (2%) in our oral cavity which are non-pathogenic for healthy individuals [9]. There is a crosstalk between the host and the microbiota, which is highly influenced by various factors like lifestyle and behaviors, tobacco consumption, oral hygiene, use of antibiotics, and the industrial revolution [10, 11]. The oral microbiota plays critical roles in protecting against colonization by extrinsic harmful bacteria [12]. Systemic diseases can affect the oral microbiome significantly [13]. It has been found that certain diseases often modify the existing microflora of the oral cavity, as shown in a study on patients of gout and hyperuricemia [12]. In this study, it was seen that the predominant microflora of the oral cavity in these group of patients as compared to healthy controls was salivary Prevotella intermedia and lower levels of Serratiamarcescens.

At present, there is no study to throw light on the alteration of oropharyngeal (OP) or nasal cavity bacterial microbiota with masks. The primary aim of the study was to see any change in Nasal and OP bacterial microbiota with the use of prolonged medical masks during the COVID-19 outbreak. The study also tried to see the prevalent antibiotic resistance pattern, as reported from sensitivity analysis reports.

## Materials and methods

Data was collected for Oropharyngeal and Nasal swabs from 30 healthy Healthcare Worker participants using N95 or Surgical Masks for at-least 80 days before the date of study initiation from June 13 to July 5, 2020. All the participants gave prior written informed consent, and all the processes of the declaration of Helsinki were followed. The study was approved by the ethical committee of Remedy Clinic Study Group (Registration Number-S/2L/59460). Participants were allowed to participate if they met the pre-defined inclusion criteria of: a) Healthcare workers using N95 masks or surgical masks for at least 8 hours a day for 80 days during COVID-19 outbreak, b) age 20 – 60 years, c) non-diabetic, d) no on-going fever during the total 40 days period before sample collection, e) no use of antibiotics in last one month, f) no halitosis and, g) no long-term use of proton pump inhibitors (more than 30 days). They were excluded from the study if: a) anyone had a history of hospitalization or fever of any cause in the last 40 days of recruitment, b) anyone developing fever by the time the culture reports are available, and c) persons not giving consent.

## Reagents and equipment

Blood agar of Biomerieux ® (Lot No: 1007843450) was used for growth and culture of fastidious organisms and Macconkey Agar of Himedia ® (Lot No: 0000411827) for growth and culture of gram-negative organisms. Swab sticks used were made of sterile cotton with polypropylene stick in high density polyethylene (HPE) tube of Himedia ® (Code No: PW009). The automated machine used for sensitivity analysis for antibiotic sensitivity was Vitek-2 ® compact.

## Microbiological procedures in nutshell

The methodology was followed as per a pre-print methodology article by the first author [14]. Nasopharyngeal swab and oropharyngeal swab from each patient were collected under aseptic precautions using a sterile cotton swab and processed immediately. Each swab was examined for both Gram’s staining and aerobic culture & sensitivity. Gram’s staining was performed to note various morphological types of bacteria in each sample. For culture, each swab was inoculated on 5% sheep blood agar (BA) and MacConkey’s agar (MA) at 37 degrees Celsius in the presence of 5-10% CO2 and, at 37 degree Celsius respectively for overnight incubation. Inoculation was done as described by *UK Standards for Microbiology Investigations Inoculation of culture media for bacteriology, n.d* [15]. The isolates grown on various cultures after overnight incubation were identified based on microscopic morphology, Gram’s staining characteristics, and automated identification system Vitek-2compact ® using standard microbiological procedure. Antimicrobial susceptibilities for aerobic bacterial isolates were performed by using Vitek-2 compact automated system ® [16], and Minimum Inhibitory Concentration (MIC) of each antimicrobial drug was reported as per the Clinical and Laboratory Standards Institute guidelines 2020 (CLSI 2020). The whole process is summarized in figure 1 below.

**Figure 1.**
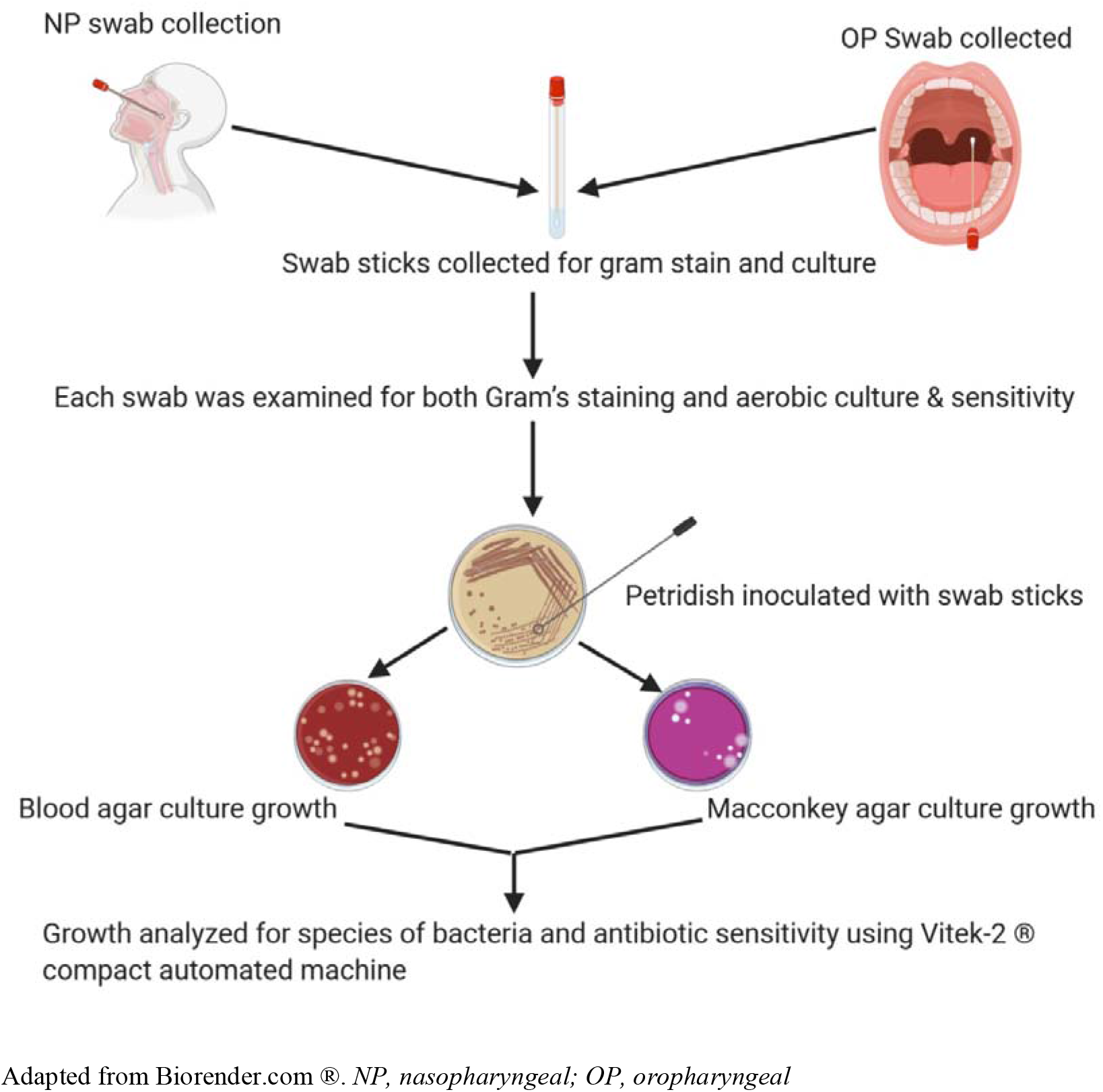
Summary of the whole microbiological processes.

## Results

The baseline characters of all the 30 participants with their standard deviation of age, BMI and weight calculated using the online calculator (https://www.calculator.net/standard-deviation-calculator.html) is depicted in table 1.

**Table 1.**
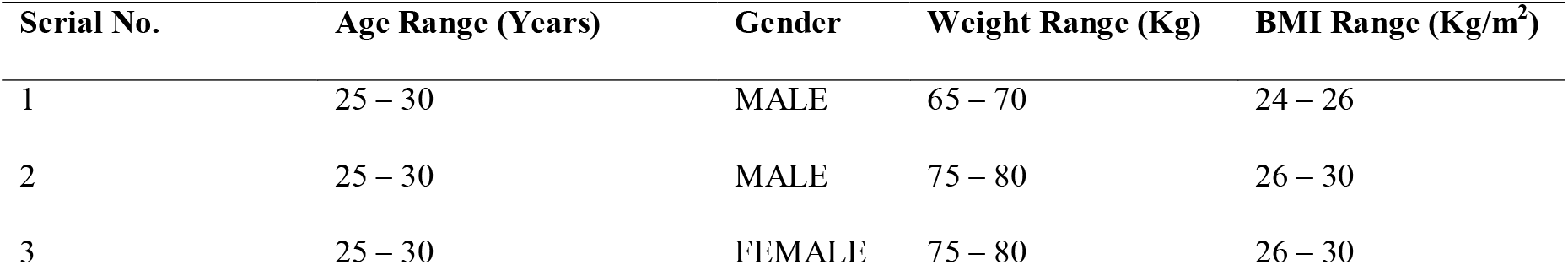
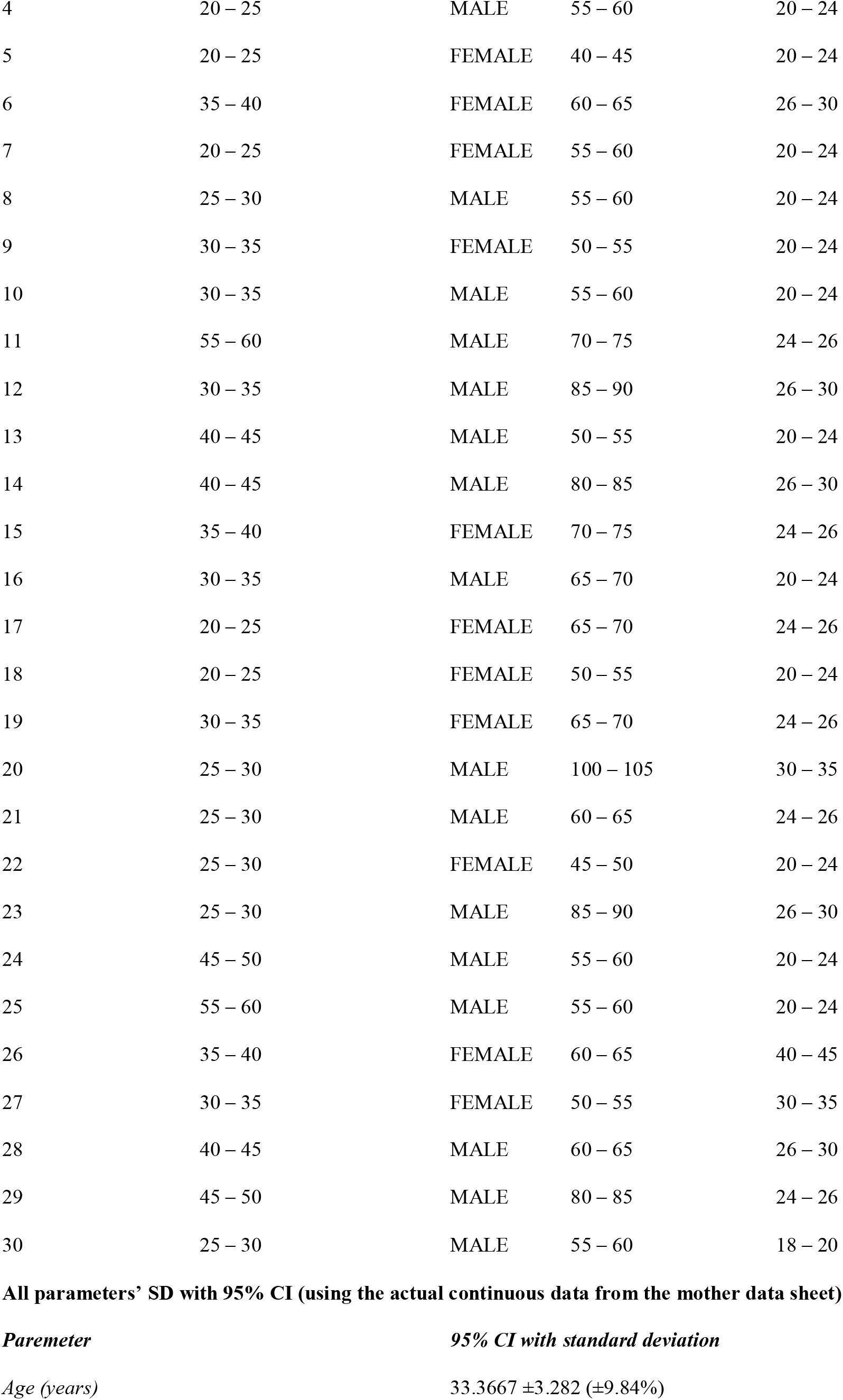
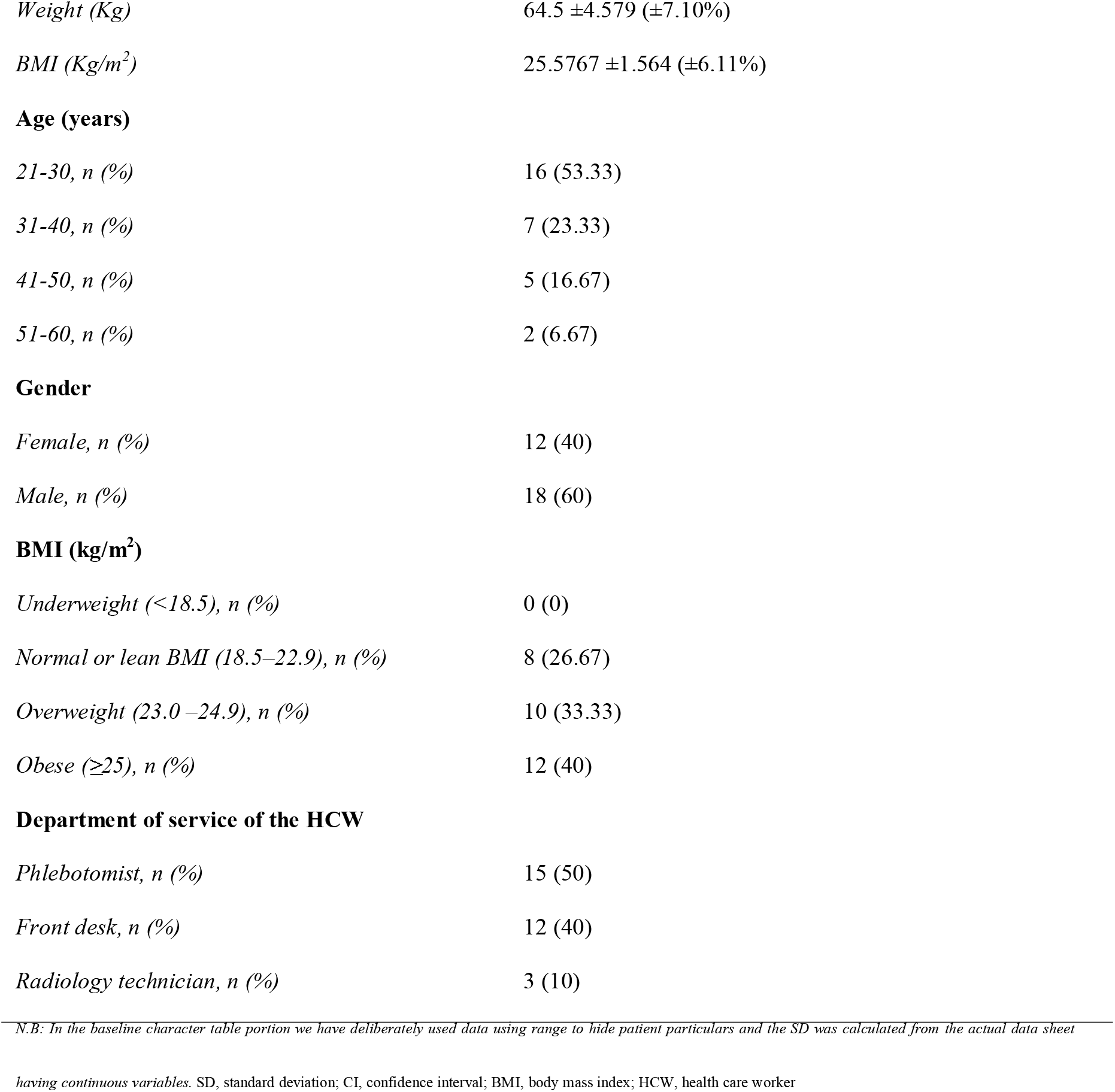
Baseline characters of the cohort (using categorical values for age, weight and BMI)

There were 10 participants showing no growth (7 in OP swab and 3 in nasal swab).Hence, 20 samples were available for final analysis which had growth both in nasal and OP swabs. The results of total thirty patients are summarised in table 2 for both nasal and OP swab cultures and gram stain findings.

**Table 2.**
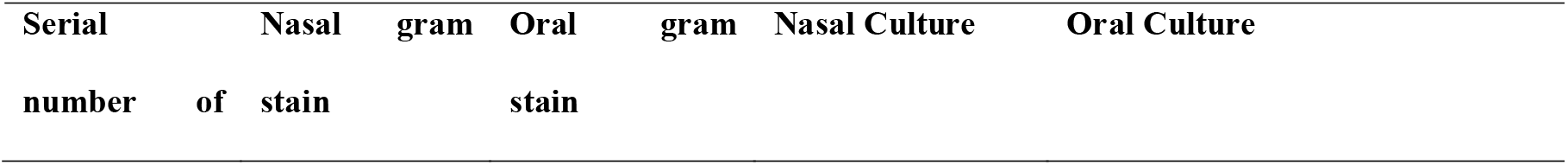
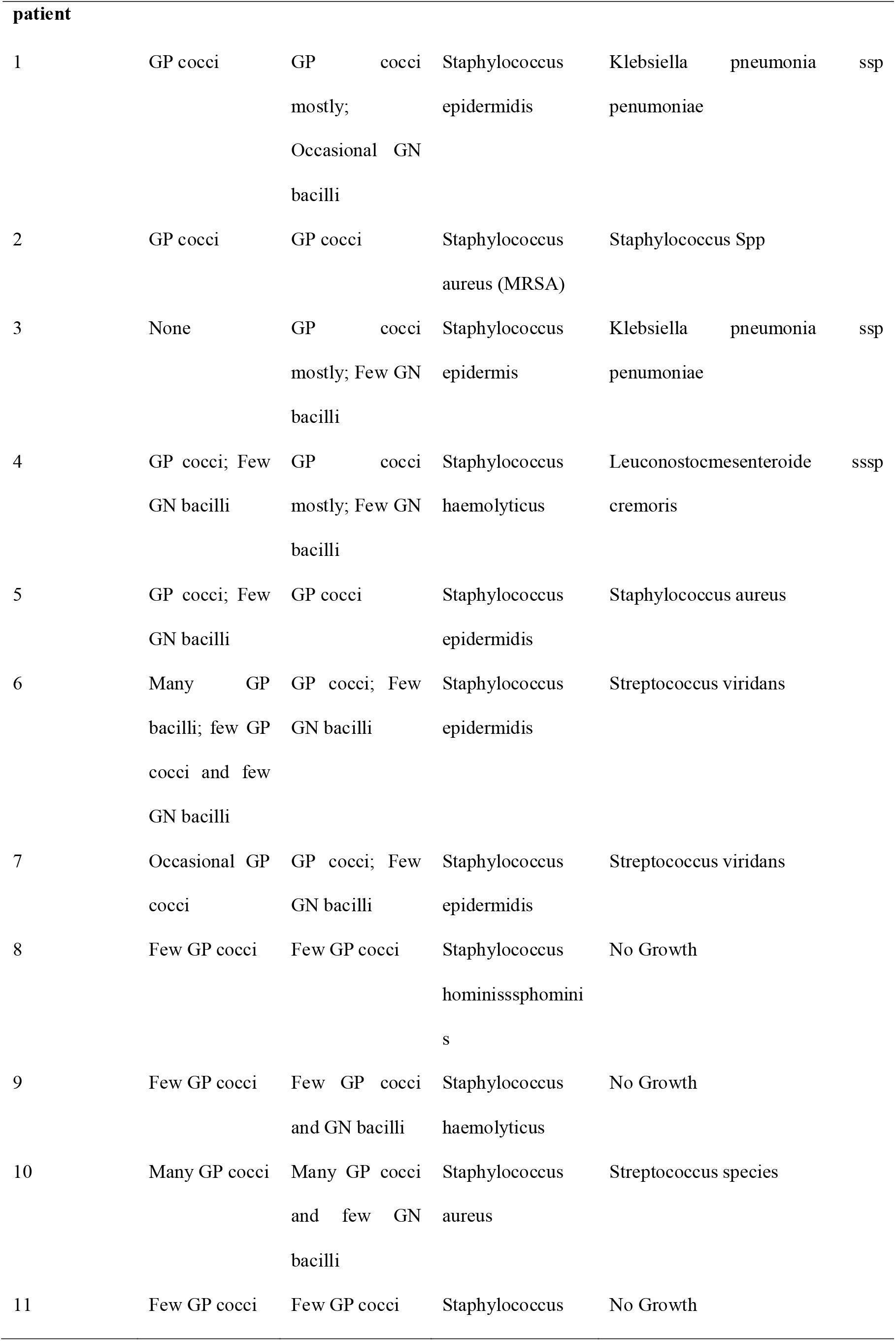
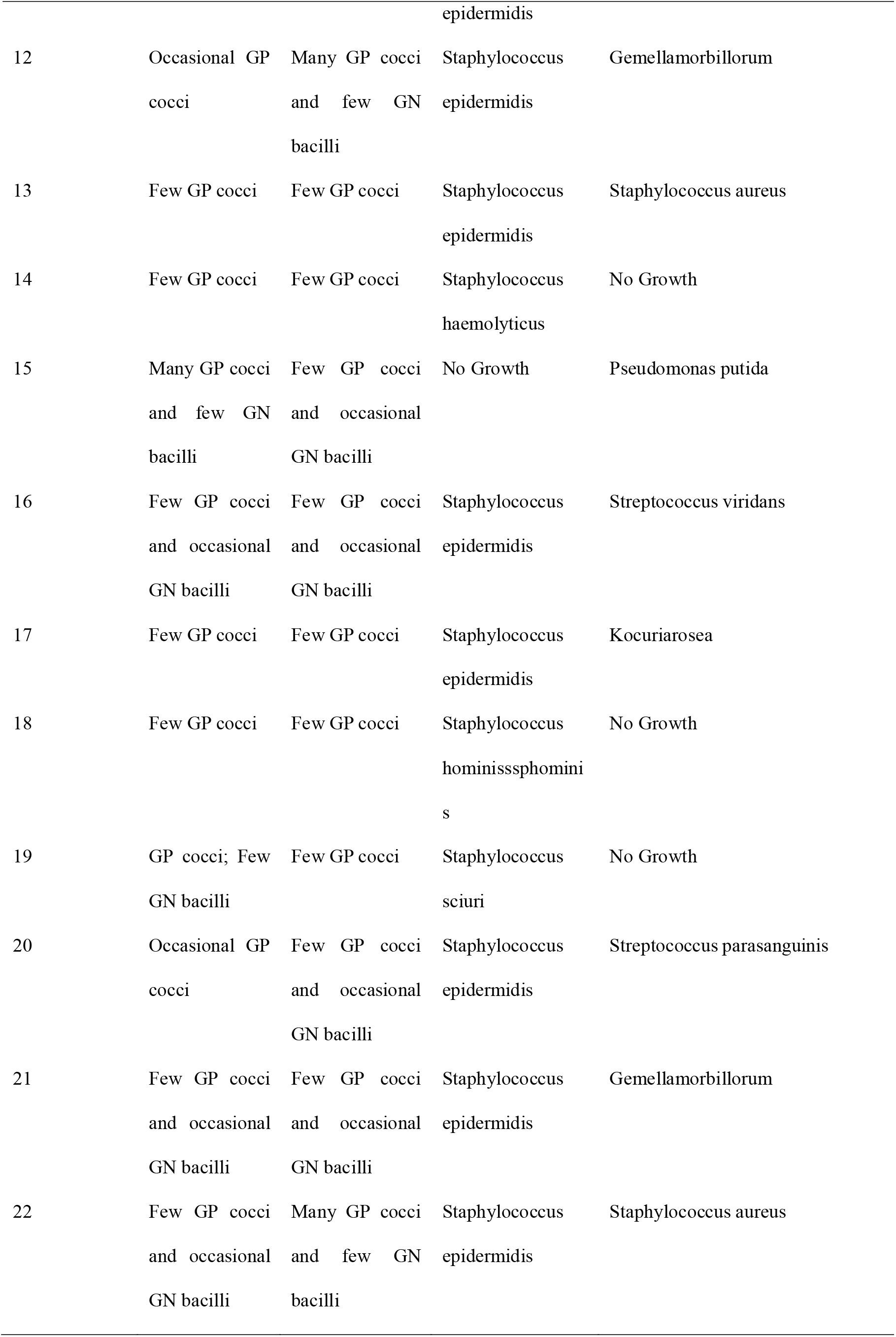
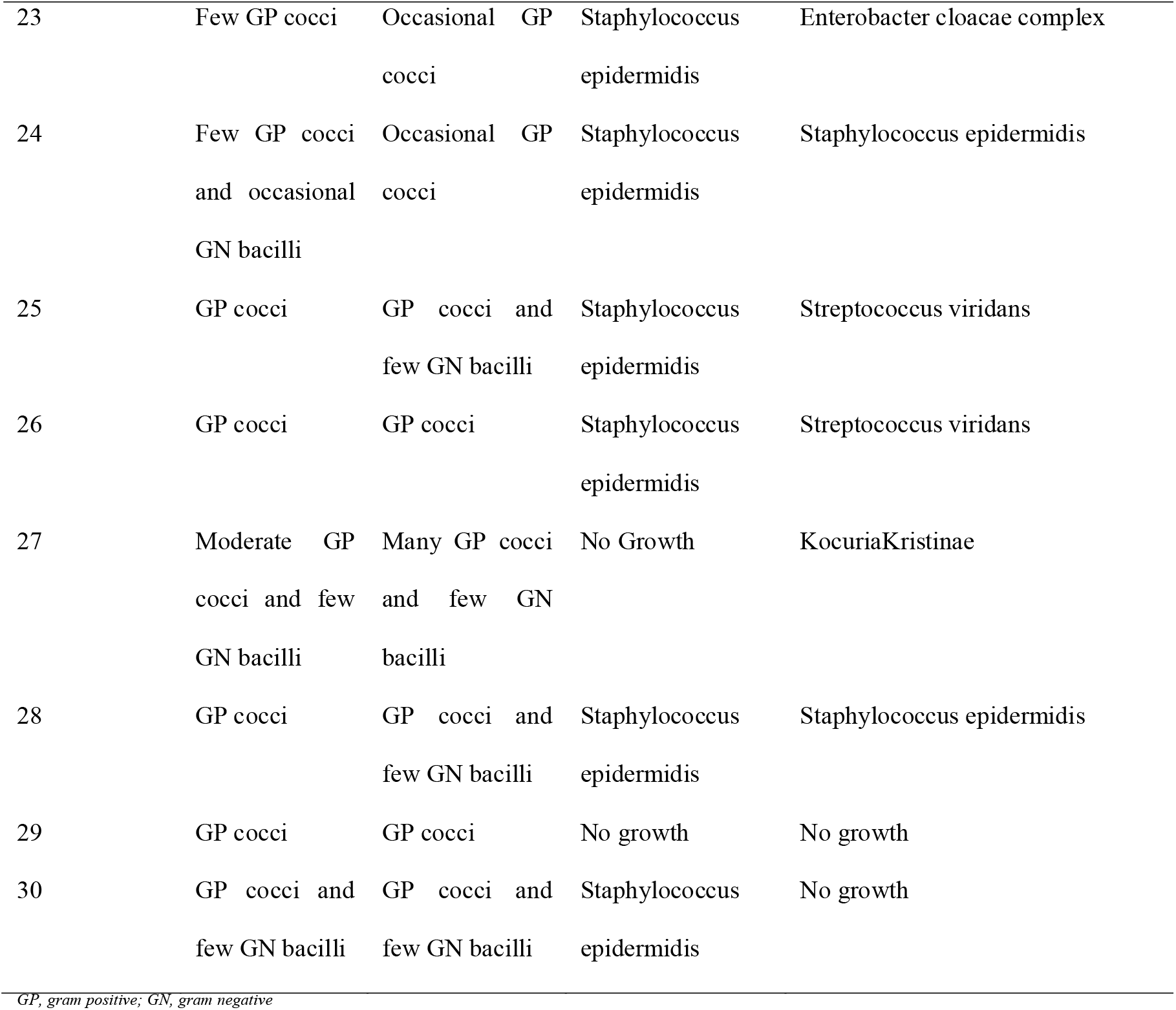
Gram stain findings and cultures of nasal and oral microflora of all participants.

The sensitivity and resistance percentage of various antibiotics of the microflora from all the participants are summarised in table 3.

**Table 3.**
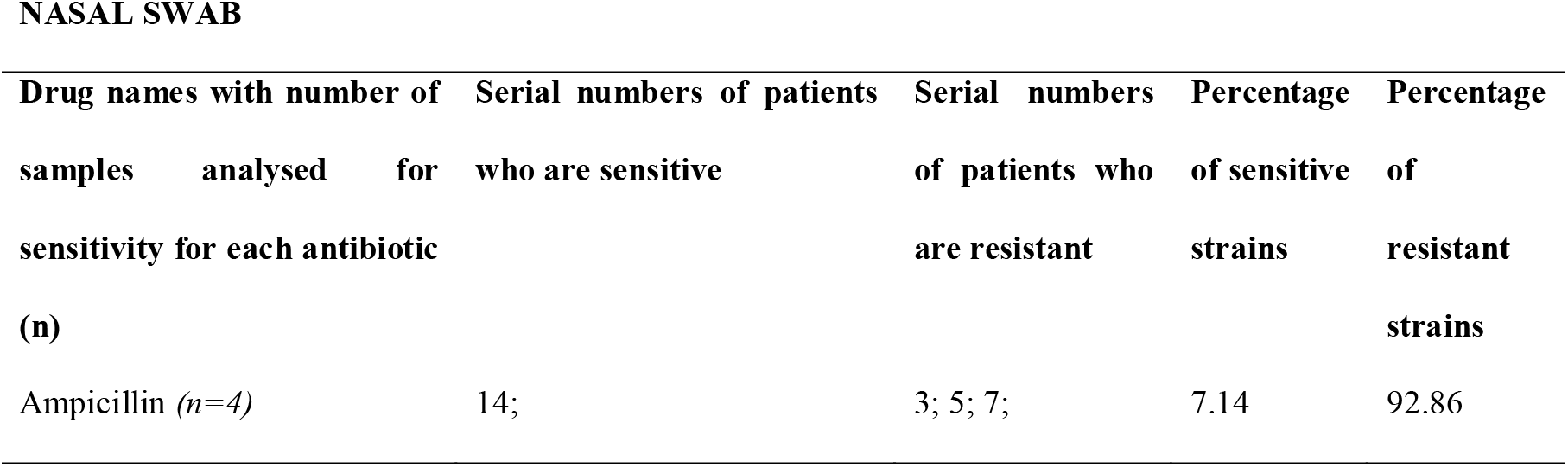
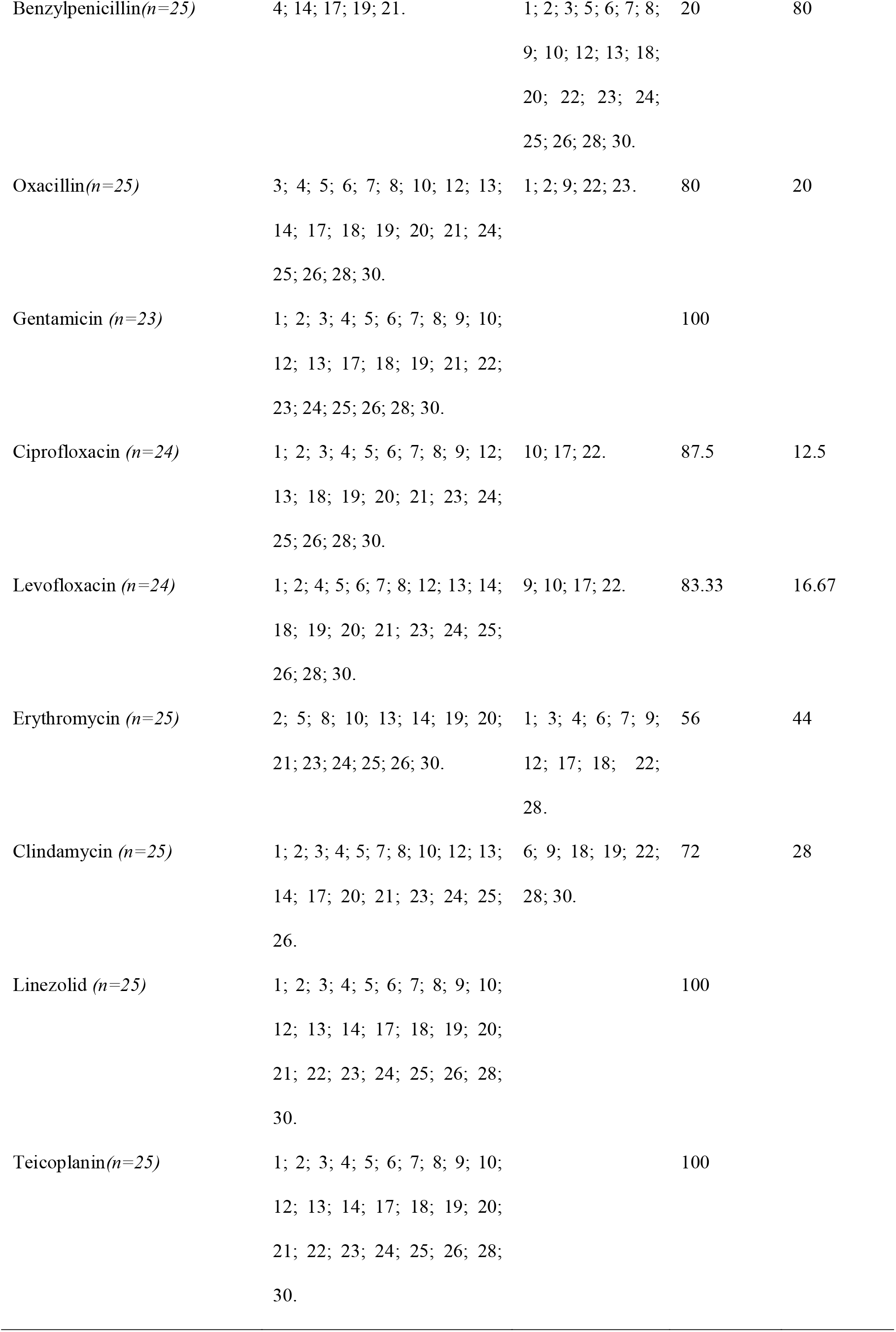
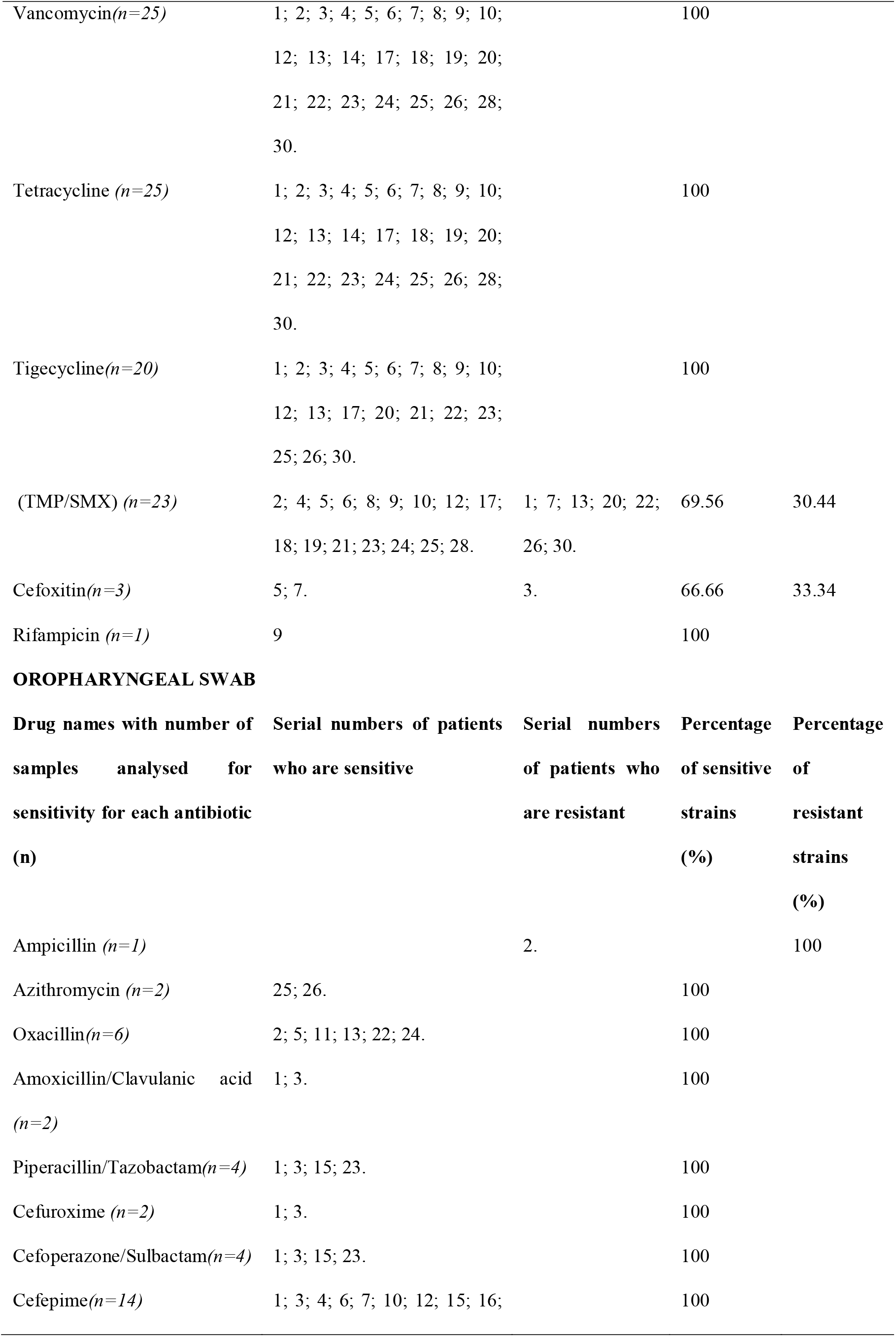
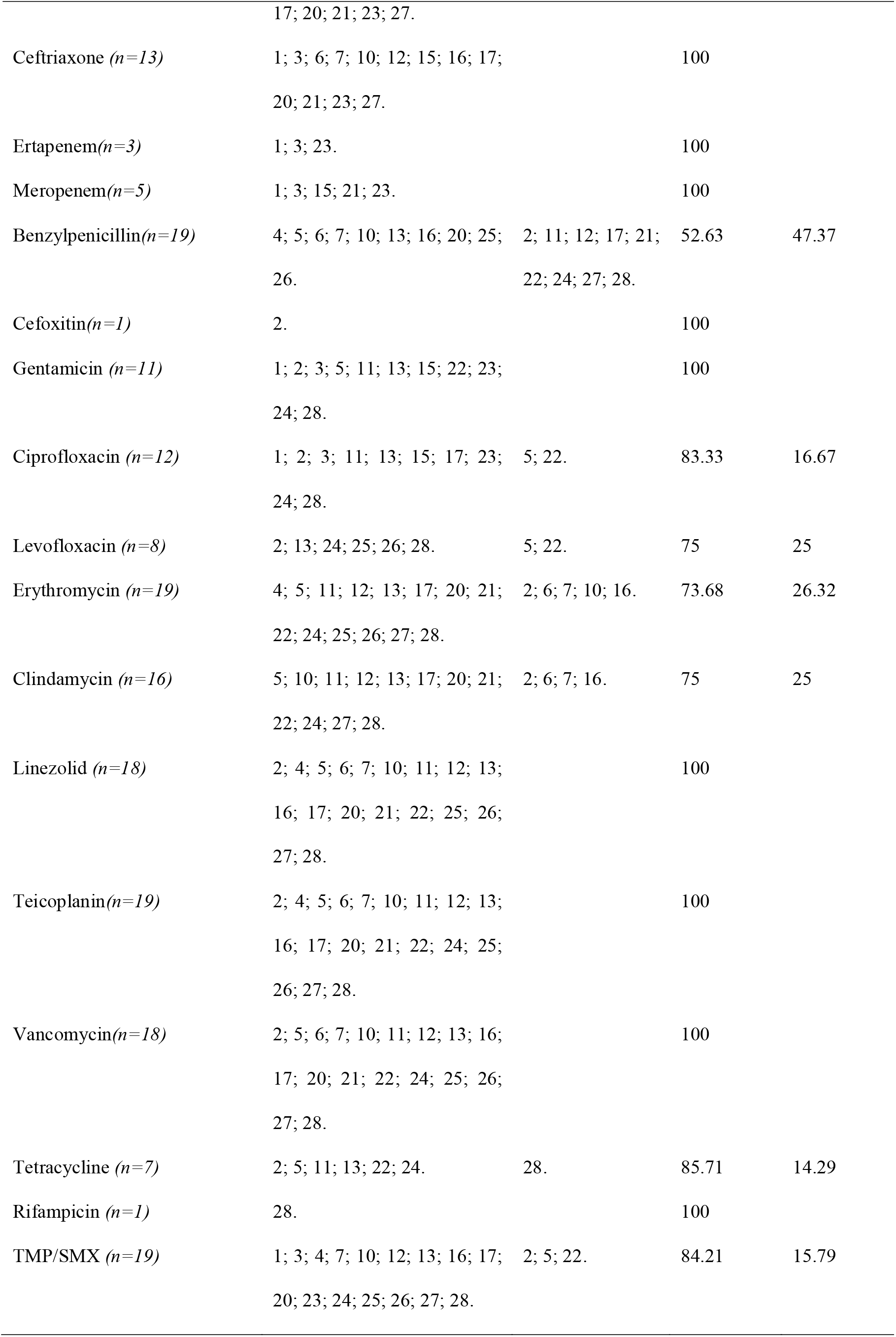
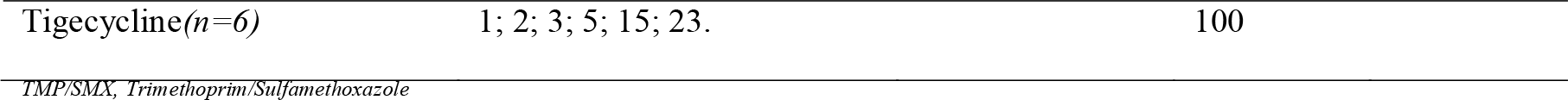
Sensitivity and resistance percentage of various antibiotics from nasal and oral swabs of all participants.

The percentages of all the species of bacteria isolated from nasal and oral cavity are given in table 4.

**Table 4.**
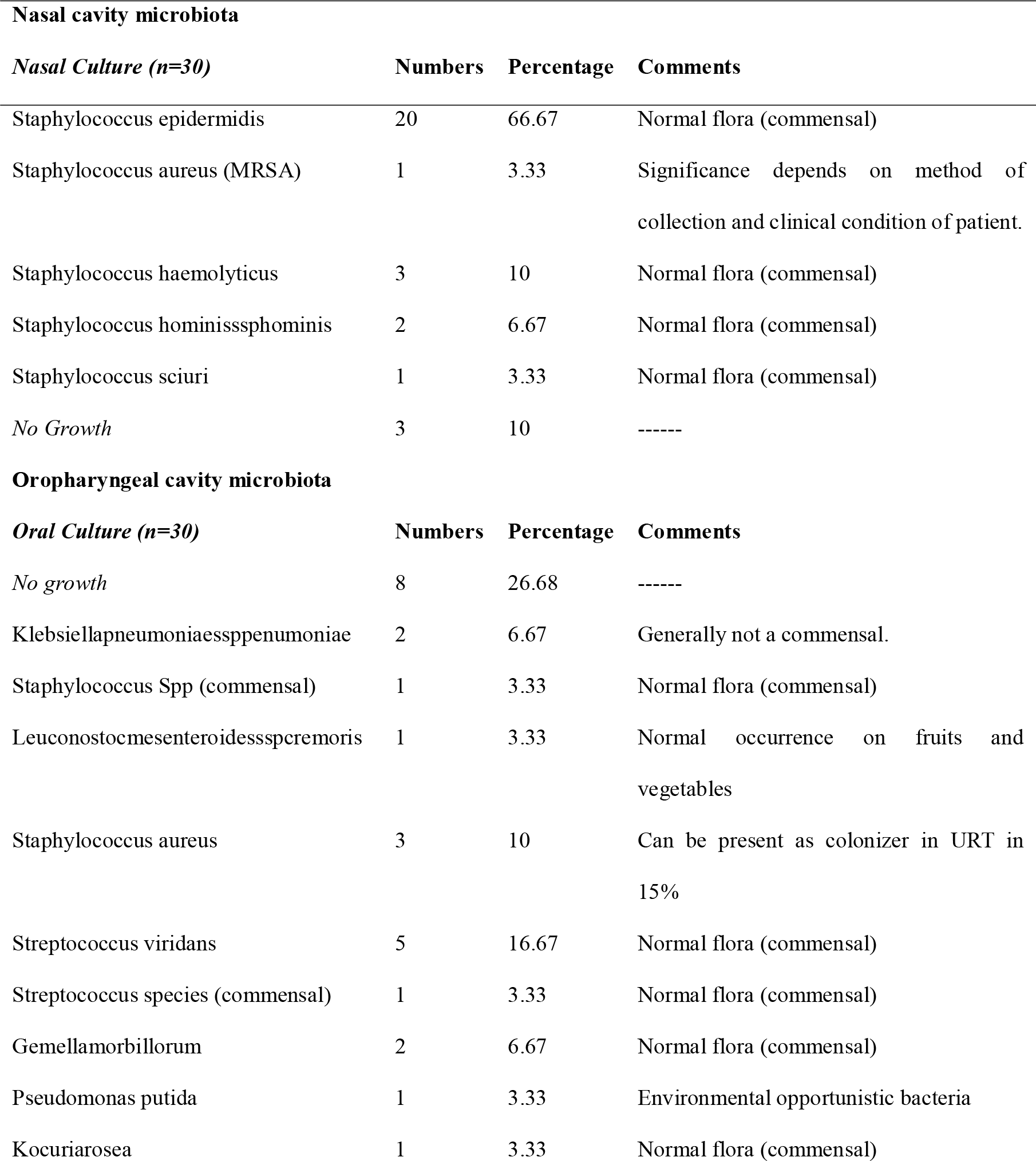
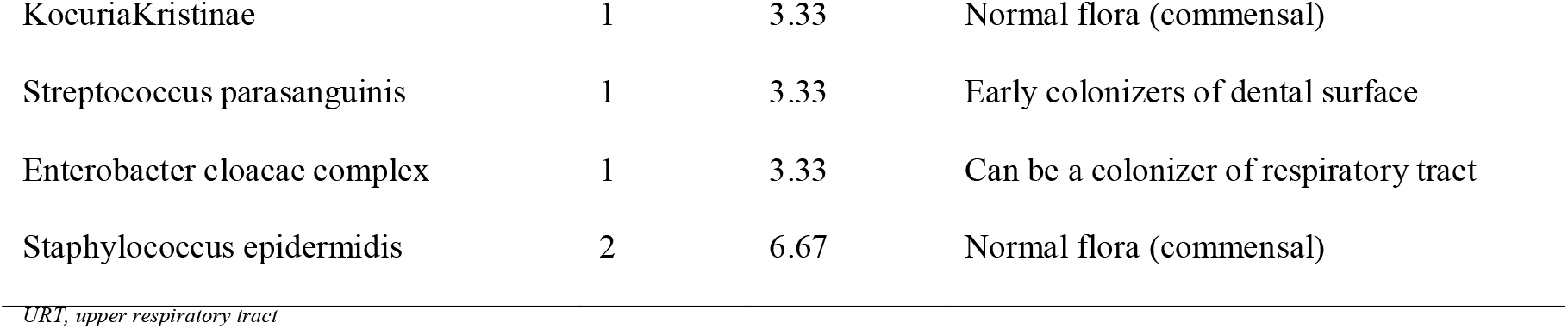
Percentages of various bacterial species in nasal and oropharyngeal cavity after using mask over prolonged period.

The participants having growth of *Staphylococcus aureus* (serial numbers 5, 10, 13 and 22) and those having growth of *Klebsiella pneumonia ssp penumoniae* (serial numbers 1 and 3) did not report any clinical symptoms suggestive of upper respiratory infections.

## Discussion

Microbiota of the body of our body varies from one compartment of the body to another, as well as significant inter-patient variability [17]. A study done to see changes in healthy nasal microflora of divers have shown an increase in nasal flora commensals with the predominant flora being *Staphylococcus epidermidis* [18]. The microflora in normal individual changes with age and lifestyle factors [19]. Up to 80% of the oropharyngeal cultivable flora is comprised of Streptococcal species, diphtheroids, and Veillonellae and Viridans streptococci predominates the nasal microflora [20].

The COVID-19 is spreading too fast to be controlled by any country and has a basic reproduction number (R0) of 2-2.5, indicating that 2-3 persons will be infected from an index patient [21]. Most healthcare set-ups and international bodies are now recommending to use various masks for prevention [22]. A survey-based study on 51 healthcare workers (HCW) highlighted the discomfort faced by HCW while wearing N95 masks [23]. The pain and discomfort while using N95 masks have also been previously raised by many other studies [24, 25]. With these views in mind during the COVID-19 outbreak, this pilot study was conducted, which showed that the normal commensals are not affected when medical masks are used over prolonged periods. The finding of no significant change is a welcome thing, since many diseases have been linked to the nasal and OP commensal microflora alterations. The study has also shown a high percentage of antibiotic resistance for Ampicillin (92.86% from nasal isolates and 100% from OP isolate), Benzylpenicillin (80% from nasal isolates and 47.37% from OP isolates) and, Erythromycin (44% from nasal isolates and 26.32% from OP isolates).

### Limitations of the study

- Non-randomized study and hence had to depend on the verbal assurance of participants
- Due to financial limitations, only bacterial flora was assessed and not any other micro-organism which are also present as commensals
- A case-control study would have been a better option, but no person was allowed to go out of home without any mask during this lockdown period of India
- Smoking was not excluded at baseline, and hence that can alter the oral as well as nasal micro-biota.

## Conclusion

Many factors have been linked to the change in oral bacterial microbiota, and this study was done to see one such factor, the use of medical masks for prolonged periods. The study found no change in normal oral bacterial microbiota, and the predominant bacterial flora was found to *Staphylococcus epidermidis* in the nasal cavity, while *Streptococcus viridans* was the commonest one from oropharyngeal microflora. This is a small pilot study, and more extensive studies are needed during and after pandemic with controls to see the actual changes.

## Data Availability

Available on request with the corresponding author.

## Acknowledgement

None to declare

## Notes

**Conflicts of interest/Competing interests:** None

### Competing Interest Statement

The authors have declared no competing interest.

### Author Declarations

Remedy Clinic Study Group (Registration Number-S/2L/59460)

